# VinDr-Mammo: A large-scale benchmark dataset for computer-aided diagnosis in full-field digital mammography

**DOI:** 10.1101/2022.03.07.22272009

**Authors:** Hieu T. Nguyen, Ha Q. Nguyen, Hieu H. Pham, Khanh Lam, Linh T. Le, Minh Dao, Van Vu

## Abstract

Mammography, or breast X-ray, is the most widely used imaging modality to detect cancer and other breast diseases. Recent studies have shown that deep learning-based computer-assisted detection and diagnosis (CADe/x) tools have been developed to support physicians and improve the accuracy of interpreting mammography. However, most published datasets of mammography are either limited on sample size or digitalized from screen-film mammography (SFM), hindering the development of CADe/x tools which are developed based on full-field digital mammography (FFDM). To overcome this challenge, we introduce VinDr-Mammo – a new benchmark dataset of FFDM for detecting and diagnosing breast cancer and other diseases in mammography. The dataset consists of 5,000 mammography exams, each of which has four standard views and is double read with disagreement (if any) being resolved by arbitration. It is created for the assessment of Breast Imaging Reporting and Data System (BI-RADS) and density at the breast level. In addition, the dataset also provides the category, location, and BI-RADS assessment of non-benign findings. We make VinDr-Mammo publicly available on https://physionet.org/ as a new imaging resource to promote advances in developing CADe/x tools for breast cancer screening.

## Background & Summary

Breast cancer is among the most prevalent cancers and accounts for the largest portion of cancer deaths, with an estimated 2.2 million new cases in 2020^1^. Treatment is most successful when breast cancer is at its early stage. Biennial screening can reduce breast cancer mortality rate by 30%^2^. Among standard imaging examinations for breast cancer screening, namely mammography, ultrasound, digital breast tomosynthesis, and magnetic resonance, mammography is the recommended modality for cancer screening^3^. Interpreting mammography for breast cancer screening is a challenging task. The recall rate of mammogram screening is around 11% with a sensitivity of 86.9%, while the cancer detection rate is 5.1 per 1,000 screens ^4^. It means that 95% of called-back cases are false-positive.

With recent advancements of learning-based algorithms for image analysis^5,6^, several works have adapted deep learning networks for mammography interpretation and showed potential to use in clinical practices^7–10^. In retrospective settings, the CAD tool as an independent reader can achieve a performance comparable to an average mammographer^9^. It can be leveraged as a decision support tool that helps enhance radiologists’ cancer detection with the reading time being unchanged^10^. Furthermore, there was evidence that shows a machine learning model developed by training on data from a specific population (UK) can generalize and perform well on another population (US)^7^.

While mammography interpretation has drawn much attention, only a few datasets are publicly available to the research community. Some of the most widely used datasets are Digital Database for Screening Mammography (DDSM)^11^, Mammographic Image Analysis Society (MIAS) dataset^12^, and INbreast^13^. Although these datasets were created with precise annotations of breast abnormalities, their sample sizes are rather limited, which might not fully leverage the potential of recent deep learning networks^14^. DDSM is prevalently used for learning-based approaches due to its sizable number of exams, with 10,480 images (2,620 exams). However, DDSM was released as a digitalized scan of screen-film mammography. At the same time, the image acquisition mode used for CAD tools in clinical practice is usually full-field digital mammography. A summary of the key characteristics of these datasets is given in Table 1.

**Table 1.** Commonly used datasets of mammography.

| Dataset | MIAS<sup>12</sup> | INBreast<sup>13</sup> | DDSM<sup>11</sup> | VinDr-Mammo (ours) |
| --- | --- | --- | --- | --- |
| **Origin** | United Kingdom | Portugal | United States | Vietnam |
| **Release year** | 1994 | 2012 | 1996 | 2022 |
| **Number of studies** | 161 | 115 | 2,620 | 5,000 |
| **Number of images** | 322 | 410 | 10,480 | 20,000 |
| **Finding types** | Mass, calcification, asymmetry, and distortion | Mass, calcification, asymmetry, and distortion | Mass and Calcification | Mass, calcification, asymmetry, distortion, and other associated features |
| **Annotation** | Circle around the finding, specified by center and radius | Contour enclosing the finding | Contour enclosing the finding | Rectangle bounding box around the finding |
| **BI-RADS assessment** | No | Yes | Yes | Yes |
| **Breast density** | Yes | Yes | Yes | Yes |
| **Mode of image acquisition** | SFM | FFDM | SFM | FFDM |

To overcome these challenges, we introduce and release the VinDr-Mammo dataset, a large-scale benchmark dataset of full-field digital mammography consisting of 5,000 four-view exams with breast-level assessment and findings annotations. Mammographies were acquired retrospectively from two primary hospitals in Vietnam, namely Hospital 108 (H108) and Hanoi Medical University Hospital (HMUH). Breast cancer assessment and density are reported following Breast Imaging Reporting and Data System^15^. Breast abnormalities that need short-term follow-up or are suspicious for malignancy are marked by bounding rectangles. Following European guideline^16^, mammography exams were independently double read. Any discordance between the two radiologists would be resolved by arbitration with the involvement of a third radiologist. To the best of our knowledge, VinDr-Mammo is currently the largest public dataset (20,000 scans) of full-field digital mammography that provides breast-level BI-RADS assessment category along with suspicious or probably benign findings that need follow-up examination. By introducing the dataset, we contribute a benchmarking imaging dataset to evaluate and compare algorithmic support systems for breast cancer screening based on FFDM.

## Methods

This study was approved by the Institutional Review Board of the HMUH and H108. All the personally identifiable information and protected health information of patients were removed. Additionally, this project did not affect clinical care at these two hospitals; hence patient consent was waived. The creation of the VinDr-Mammo dataset involves three stages: data acquisition, mammography reading, and data stratification. An overview of the data creation process is illustrated in Figure 1.

**Figure 1.**
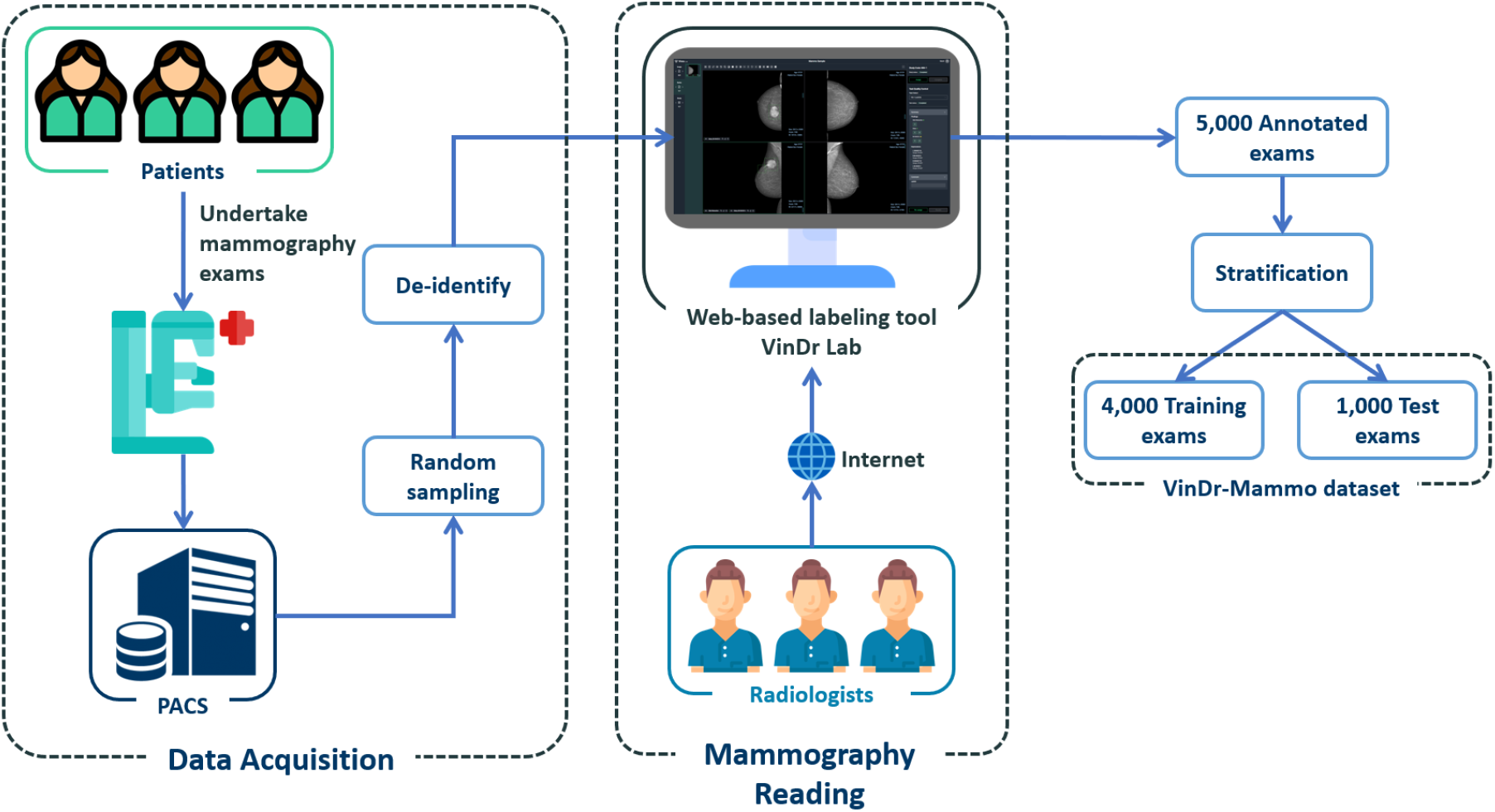
Overview of the data creation process. First, raw mammograms in DICOM format were collected retrospectively from the hospital’s PACS. These scans then got de-identified to protect patients’ privacy. Next, a web-based labeling tool called VinDr Lab was developed to store, manage, and remotely annotate DICOM data. Finally, the annotated exams were split to a training set of 4,000 exams and a test set of 1,000 exams.

### Data acquisition

In this step, 20,000 mammography images in DICOM format from 5,000 mammography examinations were sampled and collected from the pool of all mammography examinations taken between 2018 and 2020 via the Picture Archiving and Communication System (PACS) of HMUH and H108. To ensure patient privacy is protected, identifiable patient information in DICOM tags is fully removed via a Python script. Only necessary information used for loading and processing DICOM images and patient demographic information, i.e., age, is retained. Besides DICOM meta-data, associated information might appear in image pixel data, such as laterality and view position of the image and sometimes patient’s name. As this textual information usually appears in the corners of the image, we remove them by setting to black all pixels in a rectangle at each corner. The size of the rectangle is determined by visually inspecting a subset of the collected dataset. To validate the de-identification stage, both DICOM metadata and pixel data are manually reviewed by human readers.

### Mammography reading

This dataset aims to provide both the overall assessment of the breast and information of abnormal findings, which are essential to developing CADx and CADe systems for breast cancer screening. To this end, the 5,000 sampled exams containing 20,000 images were re-read, as the associated radiology reports do not indicate the exact locations of the findings.

The reading results follow the schema and lexicon of the Breast Imaging Reporting and Data System^15^. At the breast level, the overall BI-RADS assessment categories and breast density level (also termed breast composition) are provided. There are seven BI-RADS assessment categories, namely BI-RADS 0 (need additional imaging or prior examinations), BI-RADS 1 (negative), BI-RADS 2 (benign), BI-RADS 3 (probably benign), BI-RADS 4 (Suspicious), BI-RADS 5 (highly suggestive of malignancy) and BI-RADS 6 (known biopsy-proven). Since the tissue diagnosis results are not available, there is no presence of BI-RADS 6 in the re-reading process. Regarding the breast density level, its four categories are A (almost entirely fatty), B (scattered areas of fibroglandular), C (heterogeneously dense), and D (extremely dense). For the mammography findings, the list of findings provided in this dataset includes the mass, calcification, asymmetries, architectural distortion, and other associated features, namely suspicious lymph node, skin thickening, skin retraction, nipple retraction. Each finding is marked by a bounding box to localize the abnormal finding. In the given finding list, BI-RADS assessment is provided for mass, calcification, asymmetries, architectural distortion. Since the purpose of this dataset is for breast cancer screening, benign findings, i.e., findings of BI-RADS 2, are not reported to reduce the annotating time. Only findings of BI-RADS categories greater than 2, which are not confident of benign or likely to be malignant, are marked. More details of the reading reports are provided in supplementary materials. Figure 2 illustrates a sample mammography exam with both finding annotations and breast-level assessments reported by radiologists.

**Figure 2.**
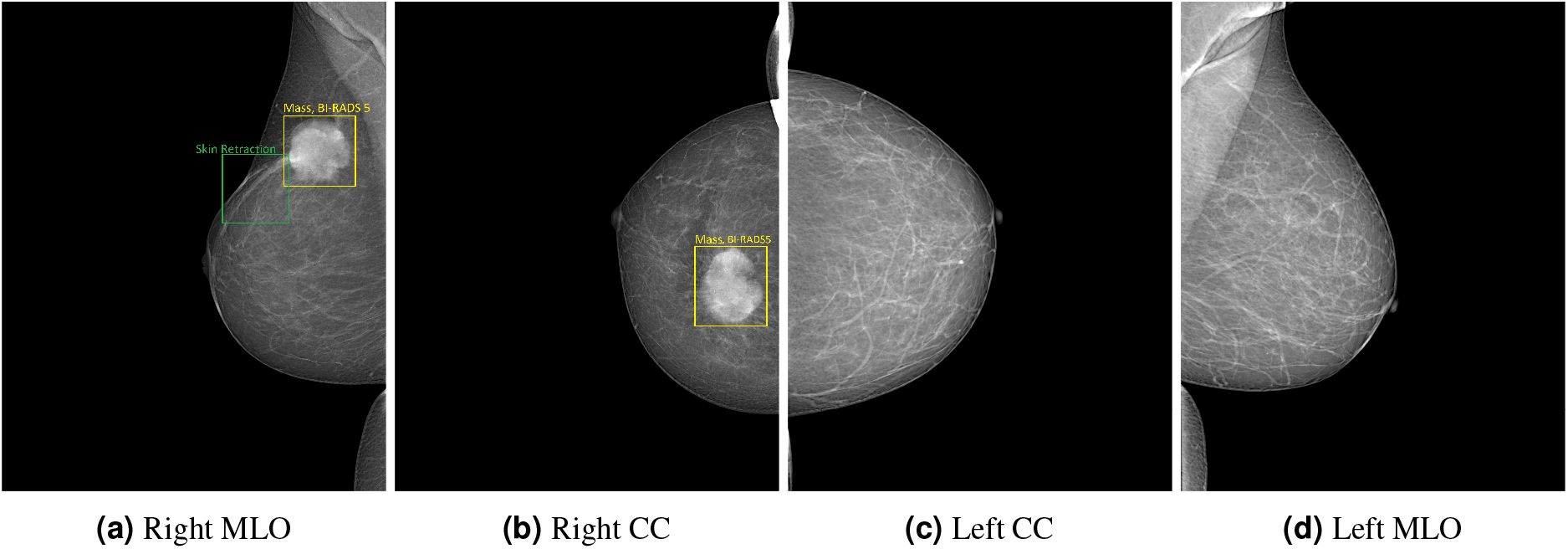
A sample mammography exam with the right breast assessed with BI-RADS 5, density B and the left breast with BI-RADS 1, density B. CC denotes craniocaudal and MLO denotes mediolateral oblique.

The mammography reading process was facilitated by a web-based annotation tool, called VinDr Lab^17^, which was specifically designed for viewing and annotating medical images. The three participating radiologists can remotely access the data for reading and annotating. All three radiologists have received healthcare profession certificates provided by the Vietnamese Ministry of Health and have more than ten years of experience. Each mammography exam was assigned to two mammographers and read independently. In cases of discordance, the exam would be assigned to the third radiologist at a higher senior experience level, to make the final decision taking into account annotations of previous readers. After the reading process had been completed, the breast level categories and finding annotations were exported in JavaScipt Object Notation (JSON) format. Subsequently, we parsed the exported file to discard unnecessary information, namely annotation timestamp, radiologist’s identifier, then simplified the file’s structure and transformed it to comma-separated values (CSV) file so that it could be easily parsed.

### Data stratification

Recent CADx and CADe solutions are mostly learning-based approaches that require separating the dataset into disjoint subsets for training and evaluation. A pre-define training/test split would help guarantee that different research works will use the same exams for training and testing. Otherwise, inconsistent or unstated splits in different research works might hinder the reproducibility and comparison of these works. For an appropriate stratification, both the training and test sets should reflex the assessment, composition, and distribution of findings of the whole dataset. However, stratifying that dataset while preserving the correlation between various data characteristics is a challenging task as the number of combinations of different attributes grows exponentially with the number of attributes (in this case are BI-RADS, breast composition, and findings categories). Hence, we split the dataset by an algorithm called iterative stratification^18^ which bases on a relaxed target that only retains the fraction of appearance of each attribute while ignoring their co-occurrence. One-fifth of the dataset, equivalent to 1,000 exams, is for testing and the rest for training. The attributes that are taken into account for splitting include breast-level BI-RADS categories, tissue composition, findings categories, and the attached BI-RADS categories (if any). The distribution of breast-level BI-RADS categories, breast composition, and findings for each subset are provided in Table 2, Table 3, and Table 4, respectively. The BI-RADS assessment of finding and patient age distribution are also depicted in Figure 3 and Figure 4.

**Table 2.** Statsitics of breast-level BI-RADS assessment.

|  | Breast BI-RADS |  |  |  |  | Total |
| --- | --- | --- | --- | --- | --- | --- |
|  | 1 | 2 | 3 | 4 | 5 |  |
| **Training** | 5,362 (67.03%) | 1,871 (23.39%) | 372 (04.65%) | 305 (03.81%) | 90 (01.12%) | 8,000 |
| **Test** | 1,341 (67.05%) | 467 (23.35%) | 93 (04.65%) | 76 (03.80%) | 23 (01.15%) | 2,000 |
| **Overall** | 6,703 (67.03%) | 2,338 (23.38%) | 465 (04.65%) | 381 (03.81%) | 113 (01.13%) | 10,000 |

**Table 3.** Statistics of breast density.

|  | Breast Density |  |  |  | Total |
| --- | --- | --- | --- | --- | --- |
|  | A | B | C | D |  |
| **Training** | 40 (00.50%) | 764 (09.55%) | 6,116 (76.45%) | 1,080 (13.50%) | 8,000 |
| **Test** | 10 (00.50%) | 190 (09.50) | 1,530 (76.50%) | 270 (13.50%) | 2,000 |
| **Overall** | 50 (00.50%) | 954 (09.54%) | 7,646 (76.46%) | 1,350 (13.50%) | 10,000 |

**Table 4.** Findings statistics on the VinDr-Mammo dataset. The number of findings and the rate of findings per 100 images are provided for the training set, test set, and the whole dataset.

| Finding | Split |  | Total |
| --- | --- | --- | --- |
|  | Training | Test |  |
| Mass | 989 (6.181) | 237 (5.925) | 1,226 (6.130) |
| Suspicious Calcification | 428 (2.675) | 115 (2.875) | 543 (2.715) |
| Asymmetry | 77 (0.481) | 20 (0.500) | 97 (0.485) |
| Focal Asymmetry | 216 (1.350) | 53 (1.325) | 269 (1.345) |
| Global Asymmetry | 20 (0.125) | 6 (0.150) | 26 (0.130) |
| Architectural Distortion | 95 (0.594) | 24 (0.600) | 119 (0.595) |
| Skin Thickening | 45 (0.281) | 12 (0.300) | 57 (0.285) |
| Skin Retraction | 15 (0.094) | 3 (0.075) | 18 (0.090) |
| Nipple Retraction | 30 (0.188) | 7 (0.175) | 37 (0.185) |
| Suspicious Lymph Node | 46 (0.288) | 11 (0.275) | 57 (0.285) |

**Figure 3.**
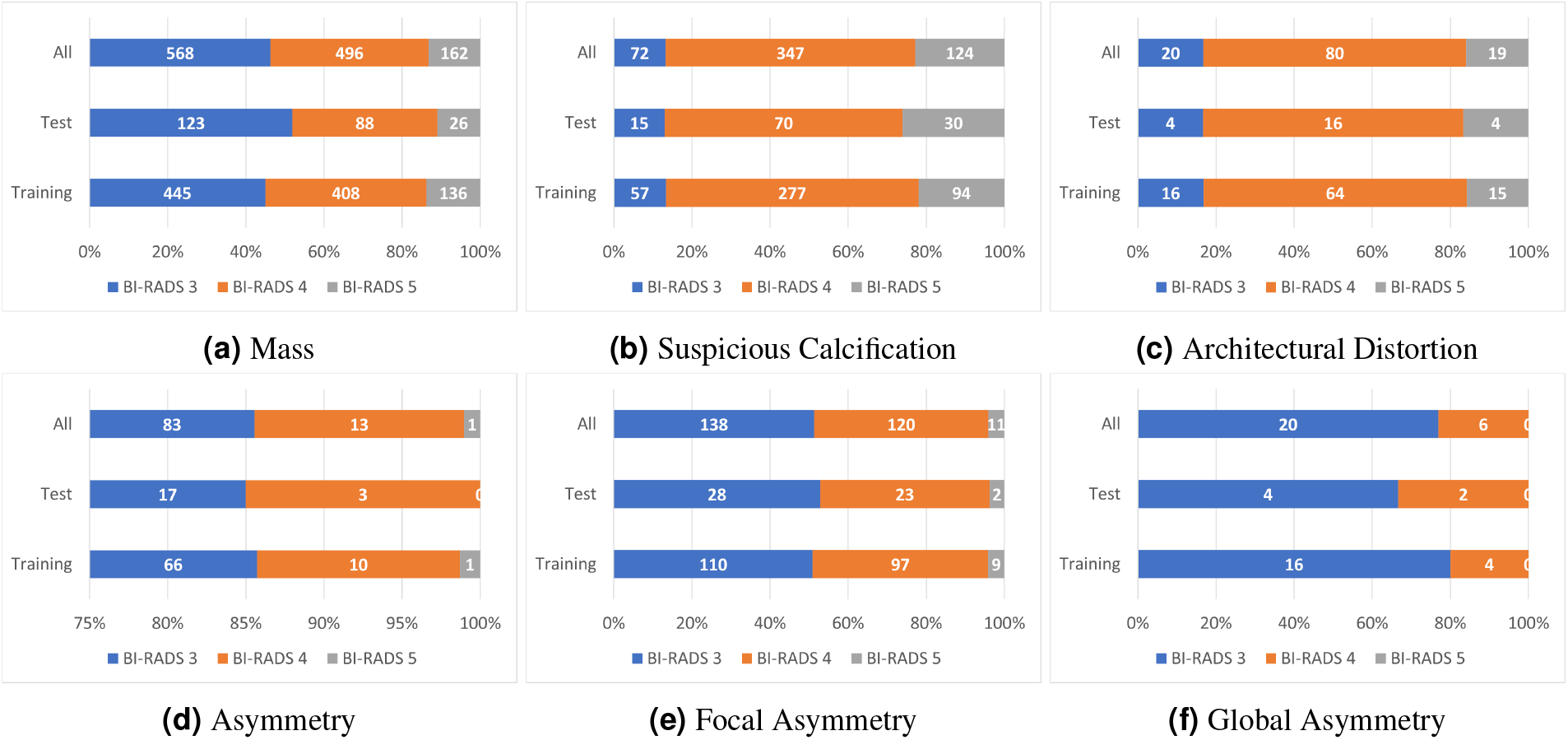
Statistics of BI-RADS assessment of findings.

**Figure 4.**
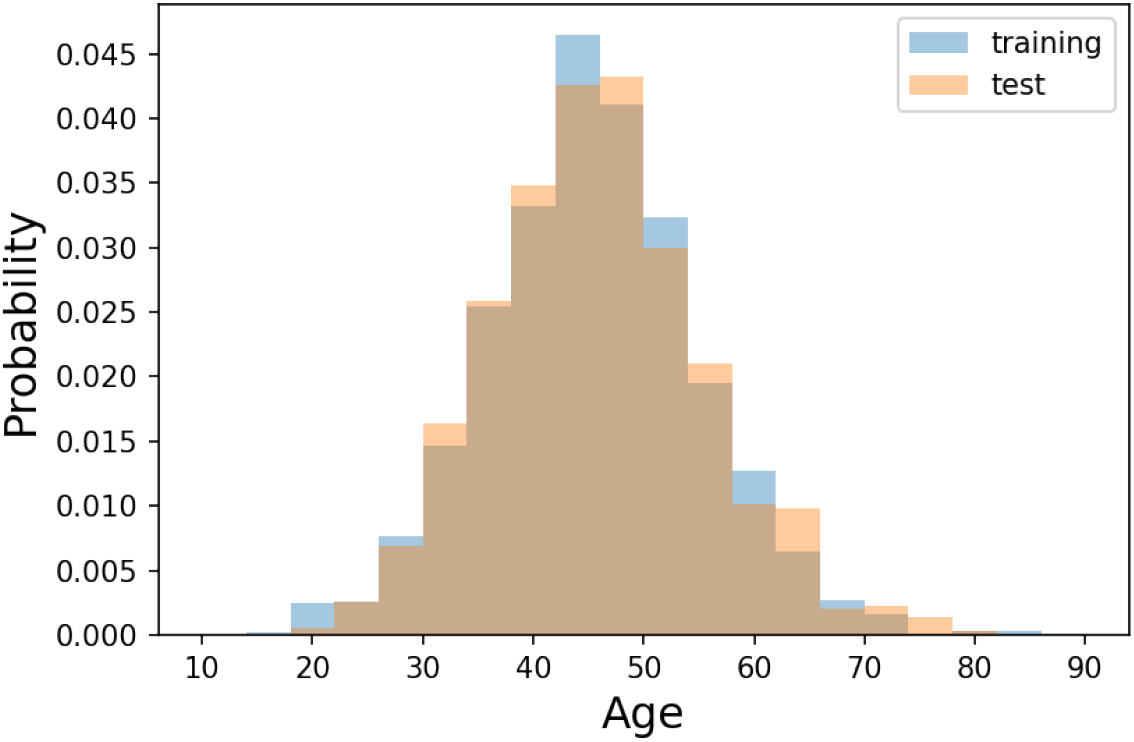
Distribution of patient age. This statistic is calculated overall all exams in which patient’s age is available.

## Data Records

Both DICOM images and radiologists’ annotations of the dataset have been submitted to PhysioNet^1^ for public access. Breast-level and finding annotations of the whole dataset are stored in CSV files `breast-level_annotations.csv` and `finding_annotations.csv`, respectively. The images are structured into subfolders according to the encoded study identifiers, each of which contains four images corresponding to four views of the exam. The subfolder name and image file name are named following the study identifier and image identifier. The information of the breast-level annotations is provided for each image even though there is redundancy since each breast is associated with two images of different view positions, i.e., MLO and CC. We find this representation more convenient because other metadata of the image, namely laterality and view position, can also be included, eliminating the need of parsing this information from the DICOM tags. Metadata for each image in the `breast-level_annotations.csv` file includes:

- `study_id`: The encoded study identifier.
- `series_id`: The encoded series identifier.
- `image_id`: The encoded image identifier.
- `laterality`: Laterality of the breast depicted in the image. Either `L` or `R`.
- `view_position`: Orientation with respect to the breast of the image. Standard views are CC and MLO.
- `height`: Height of the image.
- `width`: Width of the image.
- `breast_birads`: BI-RADS assessment of the breast that the image depicts.
- `breast_density`: Density category of the breast that the image depicts.
- `split`: Indicating the split to which the image belongs. Either `training` or `test`.

Regarding breast findings, each annotation represents the occurrence of breast abnormality at a region, represented by a bounding box, in a specific image. This means that a single finding may associate with annotations from different views, yet this linking information is not acquired in the annotation process. Metadata for each finding annotation in the `finding_annotations.csv` file contains:

- `image_id`: The encoded identifier of the image in which the finding appears.
- `study_id`: The encoded identifier of the associated study.
- `series_id`: The encoded identifier of the associated series.
- `laterality`: Laterality of the breast in which the finding appears.
- `view_position`: Orientation with respect to the breast of the image.
- `height`: Height of the image.
- `width`: Width of the image.
- `breast_birads`: BI-RADS assessment of the breast that the image depicts.
- `breast_density`: Density category of the breast that the image depicts.
- `finding_categories`: List of finding categories attached to the region, e.g., mass with skin retraction.
- `finding_birads`: BI-RADS assessment of the marked finding.
- `xmin`: Left boundary of the box.
- `ymin`: Top boundary of the box.
- `xmax`: Right boundary of the box.
- `ymax`: Bottom boundary of the box.
- `split`: Indicating the split to which the image belongs. Either `training` or `test`.

### Technical Validation

The data de-identification and the quality of the labeling process were strictly controlled. First, all meta-data was manually reviewed to ensure that all individually identifiable health information or PHI^19^ of the patients has been fully removed to meet data privacy regulations such as the U.S. HIPAA^20^ and the European GDPR^21^. In addition, pixel values of all mammograms were manually reviewed case-by-case by human readers. We developed a set of rules underlying our labeling tool to reduce mislabeling. These rules allowed to verify the radiologist-generated labels automatically. Specifically, they prevent annotators from mechanical mistakes like forgetting to choose global labels or marking lesions on the image while choosing “`BI-RADS 1`” as the breast-level assessment.

### Usage Notes

The VinDr-Mammo dataset was created for the purpose of developing and evaluating computer-aided detection and diagnosis algorithms based on full-field digital mammography. In addition, it can also be used for general tasks in computer vision, such as object detection and multiple label image classification. To download and explore this dataset, users are required to accept a Date Usage Agreement (DUA) called PhysioNet Credentialed Health Data License 1.5.0 (https://www.physionet.org/about/licenses/physionet-credentialed-health-data-license-150/). By accepting this DUA, users agree that the dataset can be used for scientific research and educational purposes only and will not attempt to re-identify any patients, institutions or hospitals. Additionally, the authors must cite this original paper for any publication that explores this dataset.

One limitation of the VinDr-Mammo dataset is that some abnormalities, namely skin retraction, and nipple retraction, have less than 40 samples, making the studies of these abnormalities on this dataset might not be reliable.

## Data Availability

All data produced in the present study are available upon reasonable request to the authors

https://physionet.org/

## Code Availability

The codes used in this study were made publicly available. The scripts used for loading and processing DICOM images are based on the following open-source repositories: Python 3.8.0 (https://www.python.org/); Pydicom 1.2.0 (https://pydicom.github.io/); and Python hashlib (https://docs.python.org/3/library/hashlib.html). The code for data de-identification and stratification was made publicly available at https://github.com/vinbigdata-medical/vindr-mammo.

## Acknowledgements

We would like to acknowledge the Hanoi Medical University Hospital and the Hospital 108 for their collaboration in creating the the VinDr-Mammo dataset and for agreeing to make it publicly available. We are especially thankful to the radiologists team Nhung Hong Luu, Minh Thi Ngoc Nguyen, Huong Thu Lai, and other collaborators who participated in the data collection and labeling process.

## Author contributions

H.Q.N. and H.H.P designed the study; H.T.N performed the data de-identification and data stratification; H.H.P and H.T.N conducted the data acquisition and analysis; H.H.P. and H.T.N wrote the paper; all authors reviewed the manuscript.

## Supplementary materials

**Table 5.** Definition of findings used in the study.

| Finding |  | Definition |
| --- | --- | --- |
| 1. Mass |  | A mass is 3-dimensional and occupies space. It has completely or partially convex-outward borders and (when radiodense) appears denser in the center than at the periphery. If a potential mass is seen only on a single projection, it should be called an asymmetry until its 3-dimensionality is confirmed. |
| 2. Suspicious Calcification |  | Calcification with suspicious morphology (amorphous, coarse heterogeneous, fine pleiomorphic, fine linear, or fine linear branching) or probably benign (BI-RADS 3). |
| Asymmetries | 3. Asymmetry | This is an area of fibroglandular-density tissue that is visible on only one mammographic projection. Most such findings represent summation artifacts, a superimposition of normal breast structures, whereas those confirmed to be real lesions (by subsequent demonstration on at least one more projection) may represent one of the other types of asymmetry or a mass. |
|  | 4. Global Asymmetry | Global asymmetry is judged relative to the corresponding area in the contralateral breast and represents a large amount of fibroglandular-density tissue over a substantial portion of the breast (at least one quadrant). There is no mass, distorted architecture, or associated suspicious calcifications. |
|  | 5. Focal Asymmetry | A focal asymmetry is judged relative to the corresponding location in the contralateral breast, and represents a relatively small amount of fibroglandular-density tissue over a confined portion of the breast (less than one quadrant). It is visible on and has a similar shape on different mammographic projections (hence, a real finding rather than superimposition of normal breast structures), but it lacks the convex-outward borders and the conspicuity of a mass. Rather, the borders of a focal asymmetry are concave-outward, and it is usually seen to be interspersed with fat. |
| 6. Architecture Distortion |  | The parenchyma is distorted with no definite mass visible. This includes thin straight lines or spiculations radiating from a point, and focal retraction, distortion, or straightening at the anterior or posterior edge of the parenchyma. |
| 7. Suspicious Lymph Node |  | Axillary lymph nodes receive lymph from vessels that drain the arm, the walls of the thorax, the breast, and the upper walls of the abdomen. Features of suspicious lymph nodes include loss or disruption of central fatty hilum, loss or pericapsular fat line, irregular outer margins, hyperattenuating, and calcified. |
| 10. Skin Thickening |  | Skin thickening may be focal or diffuse and is defined as being greater than 2 mm in thickness. This finding is of particular concern if it represents a change from previous mammography examinations. However, unilateral skin thickening is an expected finding after radiation therapy. |
| 11. Skin Retraction |  | The skin is pulled in abnormally. |
| 12. Nipple Retraction |  | The nipple is pulled in. This should not be confused with nipple inversion, which is often bilateral and which in the absence of any suspicious findings and when stable for a long period of time, is not a sign of malignancy. However, if nipple retraction is new, suspicion for underlying malignancy is increased. |

**Table 6.**
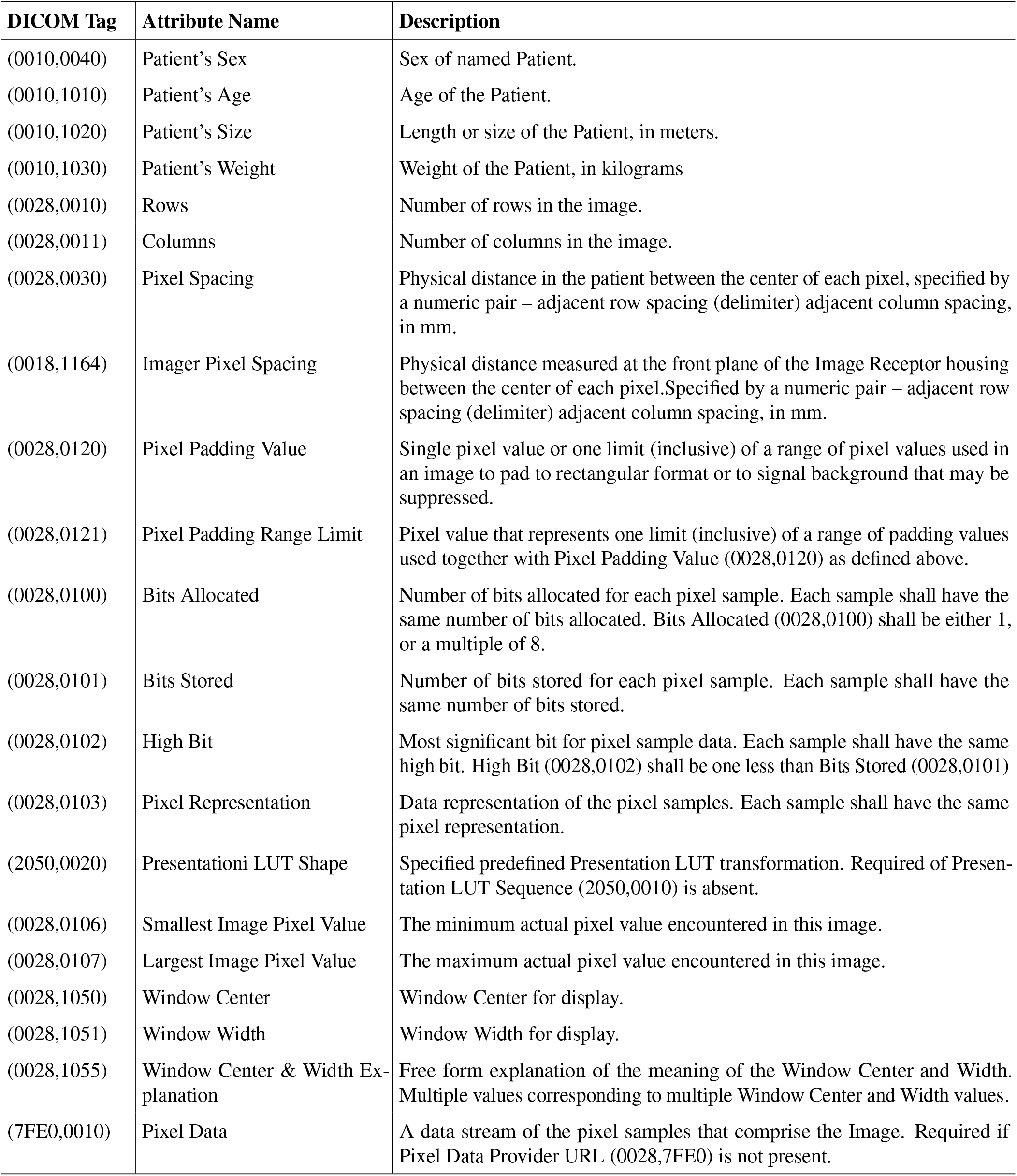
DICOM tags (a). The list of DICOM tags that were retained for loading and processing raw images. All other tags were removed for protecting patient privacy. Details about all these tags can be found from DICOM Standard Browser at https://dicom.innolitics.com/ciods.

**Table 7.**
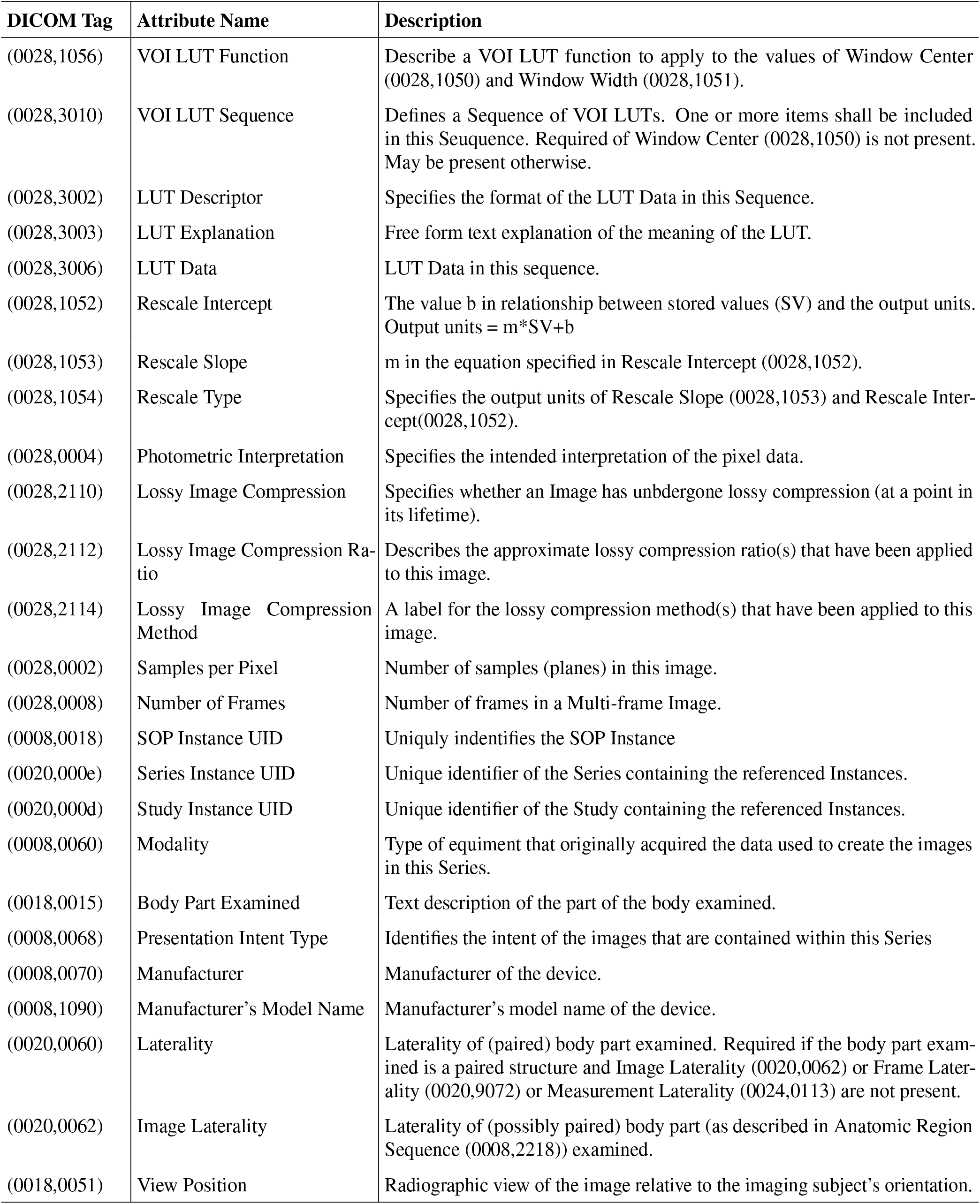
DICOM tags (b). The list of DICOM tags that were retained for loading and processing raw images. All other tags were removed for protecting patient privacy. Details about all these tags can be found from DICOM Standard Browser at https://dicom.innolitics.com/ciods.

## Footnotes

1 https://physionet.org/

